# Service availability and readiness for out-patient integrated management of multimorbidity in public primary health facilities in Malawi: A mixed methods analysis

**DOI:** 10.1101/2025.07.22.25331935

**Authors:** Gift Treighcy Banda-Mtaula, Stephen A. Spencer, Sangwani Nkhana Salimu, Ibrahim G Simiyu, Nateiya Mmeta Yongolo, Marlen S. Chawani, Augustine Choko, Paul Dark, Julian Hertz, Matthew Rubach, Francis M. Sakita, Hendry Sawe, Sarah Urasa, Adamson S. Muula, Mulinda Nyirenda, Jacob Phulusa, Ben Morton, Eve Worrall, Jamie Rylance, Miriam Taegtmeyer, Felix Limbani, Rhona Mijumbi

## Abstract

**Background:** The presence of multiple medical chronic conditions is a growing burden across Africa, where people experience a double burden of communicable and non-communicable diseases (NCD). Integrated chronic disease care at the primary healthcare level is a means to improve person-centred care for people living with multimorbidity. This study aimed to assess public primary healthcare facilities’ service availability and readiness to manage multimorbidity in Malawi, through the lens of HIV, type 2 diabetes, hypertension and chronic kidney disease. We explored healthcare workers’ perceptions of their capacity to provide care for individuals with multimorbidity and the availability of services and resources at their facility.

**Methods:** We conducted a mixed-methods health facility cross-sectional study in primary healthcare facilities in Blantyre and Chiradzulu districts, Malawi. We used the modified WHO Service Availability and Readiness Assessment Tool to assess 12 facilities, and conducted in-depth interviews with 12 purposively selected health facility leads. Availability is presented as the proportion of facilities offering each service. Facility readiness is defined as the mean(percentage) availability of 44 tracer items with a cutoff point of ≥70%, classified as ‘ready’ to manage multimorbidity. Readiness score for individual conditions was also calculated based on the availability of tracer items of that condition with a cutoff point of ≥70%.

**Results:** Of the surveyed facilities, 83%(10/12) offered services for all four selected conditions. Only 30%(3/10) of all facilities had a readiness score of ≥70%, indicative of minimum service delivery requirements for multimorbidity were met. The mean readiness score for type 2 diabetes (65.7%) was the highest, followed by HIV (63.8%), hypertension (60.4%), and chronic kidney disease had the lowest readiness score (22.9%). Barriers to service readiness included nonfunctional diagnostic equipment, frequent drug stockouts and insufficient staffing and supervision, and a lack of clinical guidelines or protocols.

**Conclusion:** The assessed sites were inadequately prepared to provide integrated care for chronic kidney disease, type 2 diabetes, HIV and hypertension. The ability to provide integrated screening and management will require functional diagnostic equipment and consistent drugs. Periodic quality assurance and combined multimorbidity audits could further highlight the gaps in service delivery and give clues to improvement.

## Introduction

The burden of multimorbidity, defined as two or more chronic medical conditions in an individual,(1, 2) is increasing in southern Africa, where there are highly prevalent communicable and non-communicable diseases.(3) Recent studies have shown a particularly high prevalence of HIV, hypertension, type 2 diabetes and chronic kidney disease (CKD),(4) with frequent co-occurrence.(5) There is also a high prevalence of overlapping risk factors such as high salt intake, obesity and hypercholesterolemia;(5, 6) and some regimens of antiretroviral therapies (ARTs) for HIV can lead to obesity, diabetes and cardiovascular disease.(7)

Primary healthcare facilities are the first point of contact for formal healthcare provision and are vital for diagnosing and managing multimorbid conditions.(8) However, delivery of quality primary healthcare in Malawi is hampered by limited resources. Existing healthcare programmes prioritise centrally and vertically delivered, single-disease care pathways, which lead to fragmented provision of care. (9) These factors impact on continuity of care for people living with multimorbidity and delay diagnosis and treatment, and can precipitate disease complications, acute hospital presentation and preventable death. (10)

Assessment of service availability and readiness within primary care is a critical first step to identify priority areas for investment and capacity building. The WHO defines service availability as the physical presence of the delivery of services (infrastructure and health personnel). Readiness is defined as the ability to deliver predetermined minimum standards that include the availability of trained staff, guidelines, diagnostic capacity and medicines and commodities.(11) These components are key markers of readiness and are central to advancing health. The WHO produced a service readiness and availability tool (SARA) to assess the availability and readiness of health facilities and to monitor and evaluate the service.(11) The SARA tool has been used to assess the readiness of facilities to provide various services, including integrated delivery of communicable diseases.(12) Integrated care models have been recommended by World Health Organization (WHO) and the Malawi Ministry of Health strategic guidelines for chronic conditions.(13, 14)

Previous availability or readiness studies for chronic conditions conducted in Malawi either focused on NCDs only or were conducted before the COVID-19 pandemic and the launch of the 2023 strategic plan, and found low availability and readiness.(15-17)

We sought to assess the service-specific availability and readiness of primary facilities to screen, diagnose and manage people living with multimorbidity as the first point of contact with the formal health sector. We defined multimorbidity as the co-occurrence of HIV, type 2 diabetes, hypertension and CKD due to their high prevalence.(4) We also explored the healthcare workers’ perceptions on education and training and capacity to diagnose and manage people living with multimorbidity.

## Methods

### Study design

We conducted a mixed-method cross-sectional study in 12 primary-level government (public) health facilities between 10 January and 8 February 2024. We collected quantitative data to determine service availability and readiness, and qualitative data to explore healthcare worker perceptions on service delivery and readiness.

This study was a nested component within the Multilink study in Malawi and Tanzania which aims to identify patients suffering from multimorbidity during emergency hospital admission, optimise treatment, and enhance post-discharge linkage to outpatient care. (10)

### Study setting and facility selection

We conducted this study in Blantyre and Chiradzulu districts in the southern region of Malawi. At the time of the study, there were 33 primary healthcare facilities in Blantyre, and 14 in Chiradzulu.

We selected study facilities at random from the World Bank’s master health facility list for Malawi (2018-2019). We only included public health centres as these provide most health care in Malawi and services are free of charge to users. We excluded specialised clinics, dispensaries and privately funded HIV and maternity units situated within primary health care. We randomly selected 8/33 facilities in Blantyre city (four in rural and four in urban strata) and 4/14 in Chiradzulu district. We purposely selected healthcare workers at the selected facilities based on their role as a facility in-charge or their delegates.

### Data Collection and Procedure

We collected data through facility surveys and in-depth interviews. We modified the WHO Service Availability and Readiness Assessment (SARA)(11) tool and the WHO Package of Essential NCDs (Additional file 1).(18) The SARA is a validated tool administered at facilities to assess the availability and readiness of the health sector to manage health conditions through assessing the presence of ‘tracer indicators’ such as functioning diagnostic equipment, common treatments and staff competency. The survey covers the availability of basic amenities, human resources and infrastructure to provide care for child health, HIV, maternal health and non-communicable diseases. For this study, we extracted sections of the survey which focused on HIV, type 2 diabetes, chronic kidney disease and hypertension.(10)The modified tool was independently verified for context suitability by one renal, one HIV and one internal medicine specialist, and suitable tracer indicators were agreed (see Additional file 2). We used a paper-based survey collection tool. GTB collected data on basic amenities at the facility, diagnostic capacity, patient care, available services, staff availability, staff training and availability of essential medicines. Where feasible, GTB directly observed the presence of absence of tracer indicators. For ease of interpretation, the scores were standardised on a 0-100 scale.

We conducted in-depth interview with the facility in-charges on the same day as readiness assessments. We used semi-structured topic guides with questions including: how they manage patients with multimorbidity; the challenges they face; and their priorities for improving care. GTB conducted the interviews in Chichewa which lasted approximately 30 minutes. Interviews additionally probed aspects of clinical reasoning and decision-making and the level of integration of services for multimorbidity.

### Data management

Completed survey data were cleaned and transferred into Microsoft excel. Interviews were audio recorded and transferred to a secure computer within 24 hours bearing the unique ID. A trained qualitative researcher transcribed and translated all interviews. Transcripts and Microsoft excel survey files were stored on a password protected computer and a Microsoft SharePoint drive which was only accessible to the Multilink study investigators.

### Data analysis

We carried out descriptive analysis for all tracer items and summarised our results with frequencies and percentages. We defined service availability as the proportion of facilities in the sample that reported offering diagnosis and management of the four pre-selected conditions individually (number of facilities offering each service/ total number of facilities x 100).

We calculated service-specific readiness for each condition, and a combined readiness for multimorbidity and a facility readiness. We defined readiness as the capacity of a facility to offer specific services through consideration of tracer items categorised into domains (Basic amenities, trained staff and guidelines, diagnostic capacity, and essential medicines, treatment and follow up).(11)

To calculate Service-specific readiness for each condition, we calculated the mean availability of tracer items for individual condition at facilities providing the service. (total number of facilities that had tracer items for conditions/total number of facilities in the survey). General readiness was calculated as a mean availability of items in each domain divided by total number of items in that domain multiplied by 100 as prescribed by the WHO SARA estimation method.(11) Facility readiness was calculated as the total number of items available divided by the total number of tracer items expected for all conditions multiplied by 100. According to SARA, facilities with a score of 70% and above meet the required level of readiness to manage multimorbidity of the preselected conditions.

GTB and FL had debriefing sessions after the first three interviews to identify key issues that needed to be further explored. GTB and FL independently coded three transcripts to compare codes. GTB, FL,RM and MT agreed on the final codebook. GTB coded and organised the transcripts using NVIVO 12 software.(19) We analysed qualitative data thematically using both deductive and inductive approach with a set of *a priori* themes to align with the domains of the survey (service delivery, health workforce, medical products, and record keeping). We considered any other emerging themes and subthemes inductively, repeatedly juxtaposing findings from the survey to triangulate the findings.

### Quality assurance

We took multiple steps along the study continuum to ensure quality. We piloted the qualitative and quantitative data collection tools to ensure contextual relevance. Where possible, GTB directly observed equipment and stock cards for survey components to validate the findings. FL and GTB iteratively reviewed the data to ensure completion and consistency. Since the interviews were conducted in the local language, GTB back-translated the interviews from English to Chichewa to maintain meaning and context. All participants validated the findings informally through listening to an interview summary, and some formally checked the context in which their quotes have been reported in our study.

## Results

We firstly present survey data and explanatory qualitative findings for each SARA domain for all conditions. We then present the service-specific readiness for each condition and the overall readiness for each facility.

### Characteristics of health facilities and study participants

Table 1 shows the characteristics of the study facilities. The facilities were located between 11km and 40km from a referral hospital. Half of the facilities (50% (6/12)) were open to patients 24 hours/day while the other half had more limited hours. The facilities had an equal distribution of male and female facility in-charges, with 66.7%(8/12) being medical assistants.(20)

**Table 1:**
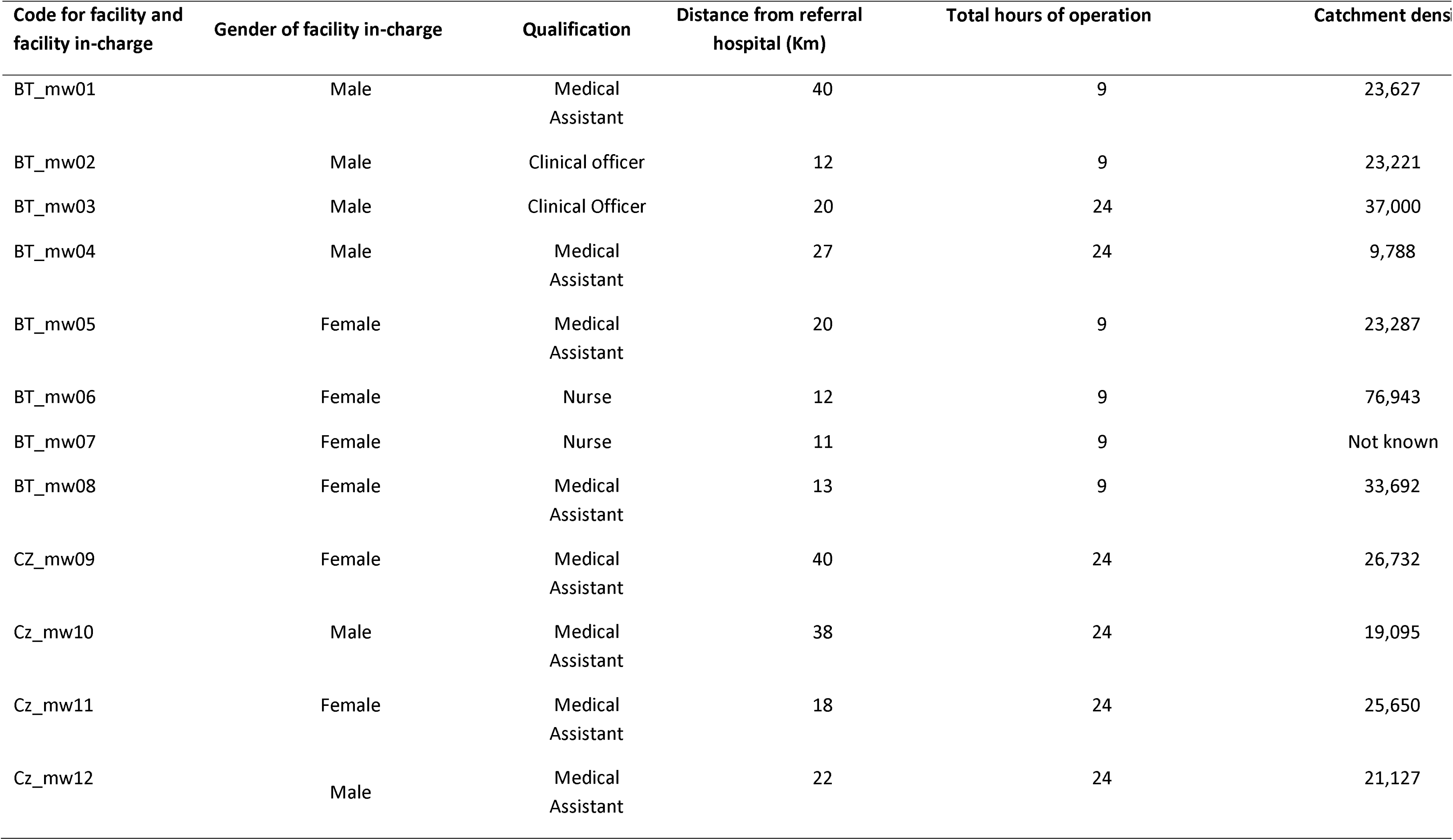
Characteristics of sampled facilities.

| Code for facility and facility in-charge | Gender of facility in-charge | Qualification | Distance from referral hospital (Km) | Total hours of operation | Catchment density |
| --- | --- | --- | --- | --- | --- |
| BT_mw01 | Male | Medical Assistant | 40 | 9 | 23,627 |
| BT_mw02 | Male | Clinical officer | 12 | 9 | 23,221 |
| BT_mw03 | Male | Clinical Officer | 20 | 24 | 37,000 |
| BT_mw04 | Male | Medical Assistant | 27 | 24 | 9,788 |
| BT_mw05 | Female | Medical Assistant | 20 | 9 | 23,287 |
| BT_mw06 | Female | Nurse | 12 | 9 | 76,943 |
| BT_mw07 | Female | Nurse | 11 | 9 | Not known |
| BT_mw08 | Female | Medical Assistant | 13 | 9 | 33,692 |
| CZ_mw09 | Female | Medical Assistant | 40 | 24 | 26,732 |
| Cz_mw10 | Male | Medical Assistant | 38 | 24 | 19,095 |
| Cz_mw11 | Female | Medical Assistant | 18 | 24 | 25,650 |
| Cz_mw12 | Male | Medical Assistant | 22 | 24 | 21,127 |

All facilities reported offering diagnosis and management of diabetes, hypertension and chronic kidney disease, and two facilities did not offer HIV services. There was substantial variation in the availability of resources for these conditions (Figure1). For service-specific readiness, CKD scored the lowest, and HIV had the highest score for diagnostic equipment, trained staff and guidelines and first line ARTs.

### Readiness to provide integrated care for chronic kidney disease, type 2 diabetes, HIV and hypertension

Only 30%(3/10) of facilities that reported offering services for all four conditions were classified as “ready” according to our pre-determined composite indicator, with not much difference between facilities open 9 and 24 hours (Additional file 3). Figure 2 shows the domain score for the integrated management of multimorbidity. Overall, diagnostic equipment and essential medicines were the lowest-scoring domains.

**Figure 1:**
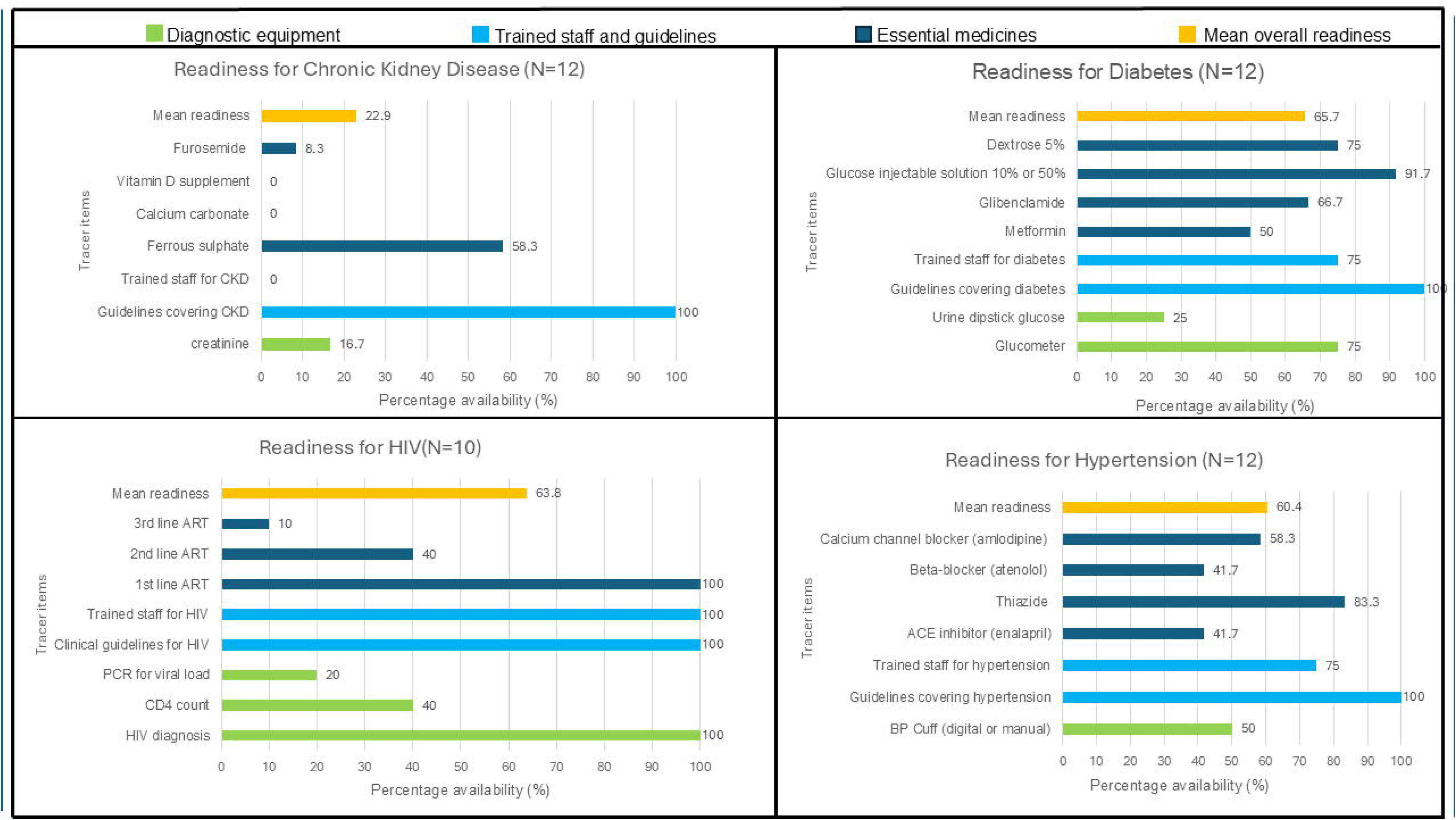
Service-specific readiness for individual conditions.

**Figure 2:**
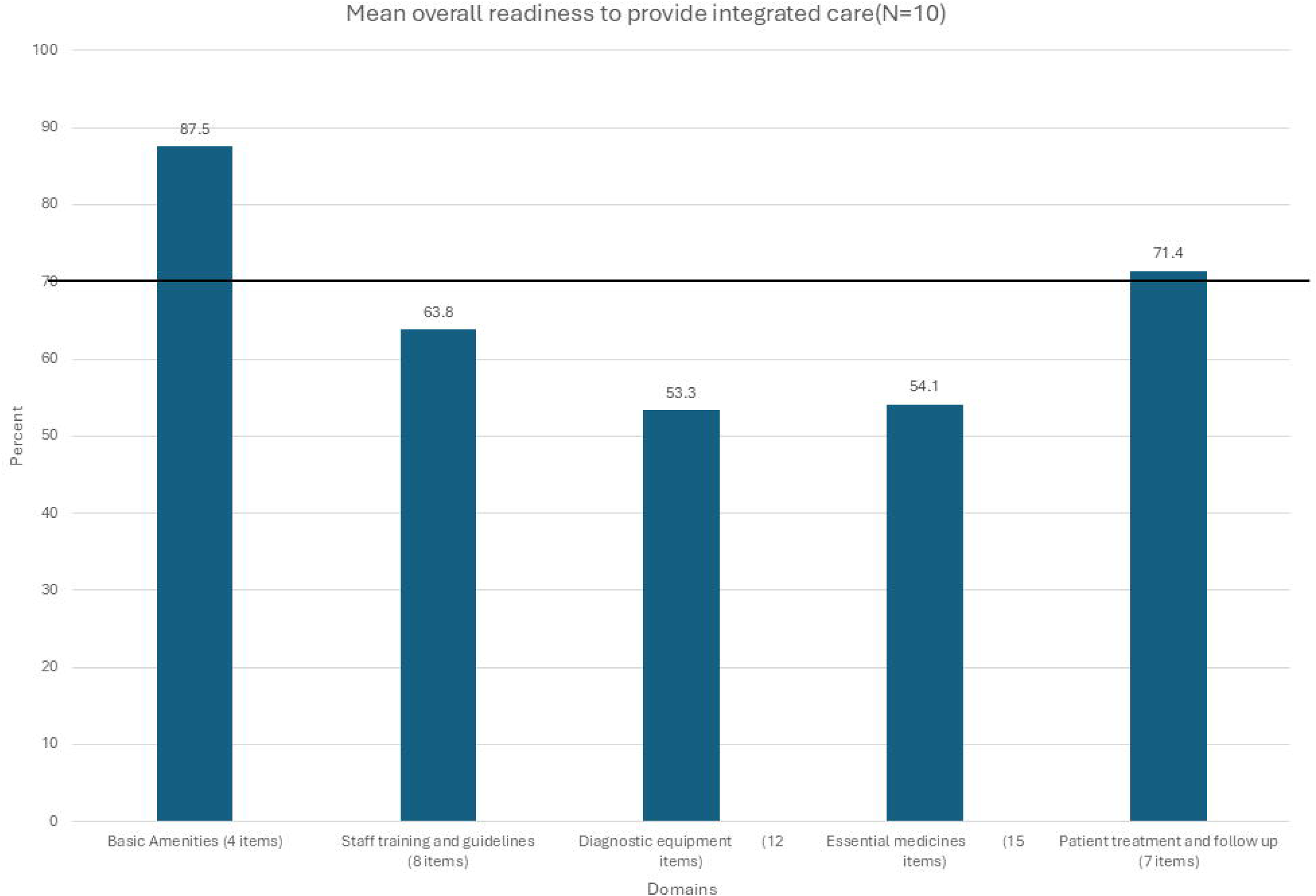
Percentage score of five domains to provide multimorbidity care. The horizontal line shows the cutoff point (70%) below which was considered “not ready”.

### Availabili

All facilities had a room for visual and auditory privacy that was used for patient consultations. However, the rooms were few and used for both acute and chronic disease consultations, giving priority to acute conditions. Sometimes patients who present with chronic conditions met **“***under a tree or on the veranda”.* The lack of a designated area for chronic conditions is seen as an inconvenience, as the outpatient department (OPD) waiting area gets congested.

Half of the facilities had a hospital mobile phone to communicate with referral facilities and other staff, while others reported a lack of phone hindered communication among staff and between staff and patients. Most facilities(7/10) had a functional computer for electronic data entry, but paper-based data collection and storage were common. All facilities also had access to an ambulance through the District Health Office (DHO), and two had extra access to an ambulance donated and managed by the area member of parliament. Ambulances were not readily available for emergency cases requiring referral to higher facilities due to challenges such as a lack of fuel.

*“On the ambulance, sometimes they tell us there is no fuel, sometimes the ambulance comes.” (BT_mw01,male,medical assistant)*

*“it’s just one ambulance that is just making rounds on first come first serve. If they have gone to A, you might hear that they will proceed to B and from there they will go to the district hospital and after that then they will come here…so that means the time is also going” (CZ_mw09,female,medical assistant)*.

### Availability of trained staff and clinical guidelines

All our participants reported at least one facility-based healthcare worker at their facility received on-the-job HIV and NCD training. All clinicians reported that all community-based healthcare workers (Health surveillance assistants) received formal HIV training and orientations to the 2022 HIV guidelines(Clinical Management of HIV in Children and Adults), with at least 3 community-based healthcare workers receiving training on NCD screening. The training was not uniform across cadres.

*“None of us (clinicians) received ART training, but all but one nurse got trained. In terms of NCDs, only a few HSAs (*health surveillance assistants*) were trained.” (*BT_mw03,male,clinical officer)

Participants reported trained HSAs assist with outpatient clinics, but are not enough, which leads to burnout.

*“During clinics, they (HSAs) give health promotion talks, and check blood pressure, but they are just a few, so rotation becomes hectic because it’s the same people who keep on coming now and again. Sometimes they lie about being sick because they are tired.”* (BT_mw07,female,nurse)”

Four facilities (4/10) reported having scheduled continuous professional development aimed at in-service and refresher training for staff members. The CPDs were reported to be inadequate to cover some of the conditions.

*“We can be doing a lot of NCDs training and not just focusing on the hypertension. We should at least look into diabetes and HIV as well. We learnt most of the things way back, we just remember here and there” (*BT_mw08,female,medical assistant)

There was varying availability of guidelines among survey facilities. Malawi Integrated Clinical HIV Guidelines were available at all the facilities that reported to offer HIV services, 60.0% (6/10) did not have the Malawi Standard Treatment Guidelines which covers the management of diabetes, hypertension and CKD individually. Similarly, 70% (7/10) did not have any wall charts or posters on prevention or management of NCDs.

### Availability of diagnostic equipment and lab services

Out of all the facilities that reported managing all four conditions, 80.0% (8/10) reported offering diagnostic capacity for blood glucose monitoring and 50% (5/10) had functional equipment for blood pressure measurement (Table 1). The equipment that was not available were either due to stock-out of consumables.

*“Sometimes our glucometer does not have glucose-sticks.” (BT_mw07,female,nurse)*

Participants reported referring patients to nearby primary facilities or higher facilities in cases where diagnostic equipment or services were unavailable. Some facilities referred patients to nearby private fee-for service health facilities.

### Availability of essential medicines

We grouped drug availability as “consistent stock within the last three months”, “inconsistent stock within the last three months”, and “never available”. All (n=10) facilities with HIV services had a consistent stock of 1^st^ line ART, and only one had first, second and third-line treatments available(Table 3). Metformin, an essential drug for type 2 diabetes, was found in half of the facilities. Most of our participants reported ferrous sulphate was often available but prioritised for maternal health and not the anaemia found in kidney disease, and most of the CKD medication were never available.

**Table 2:**
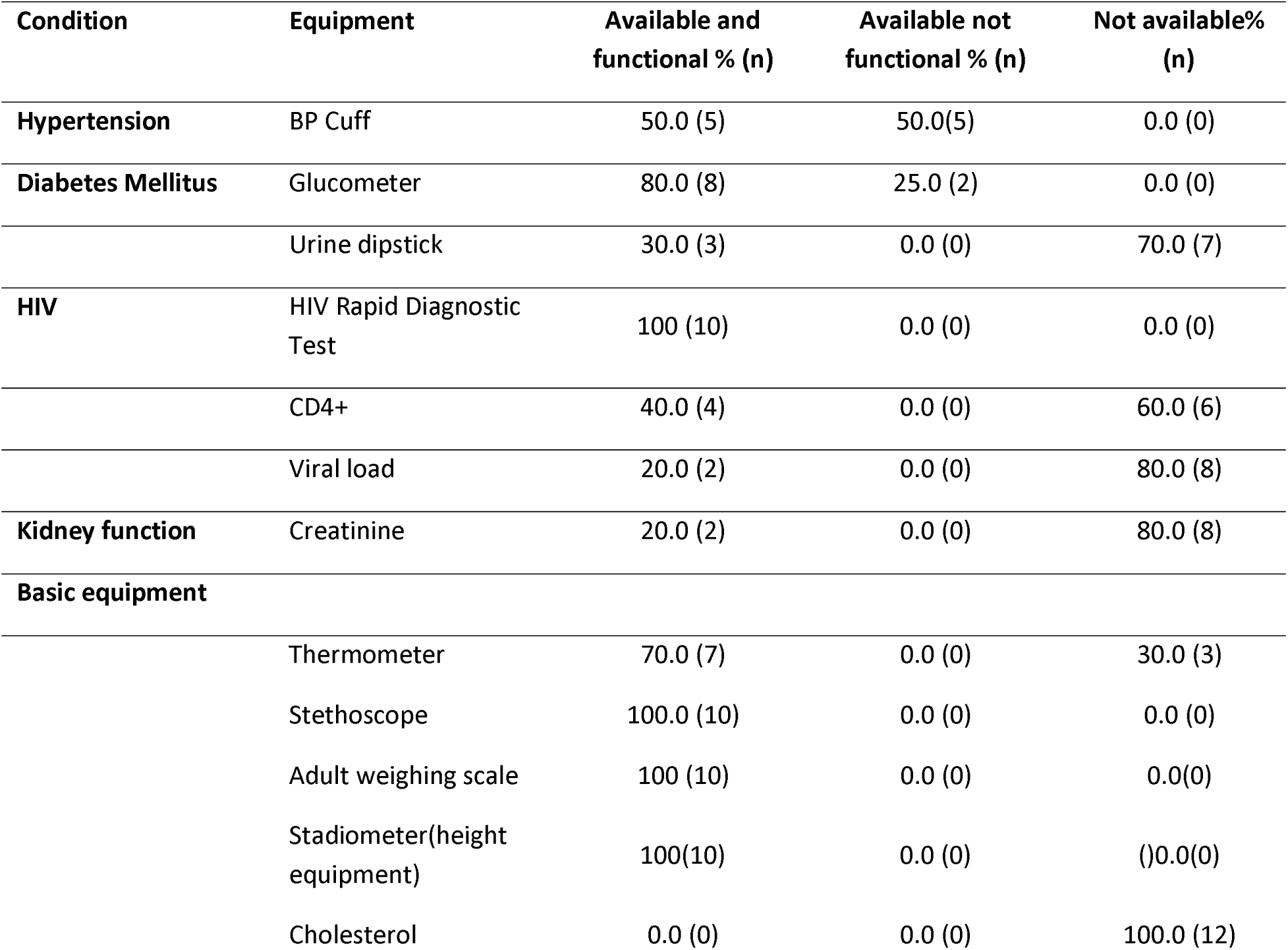
Availability of diagnostic equipment(N=10)

| Condition | Equipment | Available and functional % (n) | Available not functional % (n) | Not available% (n) |
| --- | --- | --- | --- | --- |
| **Hypertension** | BP Cuff | 50.0 (5) | 50.0(5) | 0.0 (0) |
| **Diabetes Mellitus** | Glucometer | 80.0 (8) | 25.0 (2) | 0.0 (0) |
|  | Urine dipstick | 30.0 (3) | 0.0 (0) | 70.0 (7) |
| **HIV** | HIV Rapid Diagnostic Test | 100 (10) | 0.0 (0) | 0.0 (0) |
|  | CD4+ | 40.0 (4) | 0.0 (0) | 60.0 (6) |
|  | Viral load | 20.0 (2) | 0.0 (0) | 80.0 (8) |
| **Kidney function** | Creatinine | 20.0 (2) | 0.0 (0) | 80.0 (8) |
| **Basic equipment** |  |  |  |  |
|  | Thermometer | 70.0 (7) | 0.0 (0) | 30.0 (3) |
|  | Stethoscope | 100.0 (10) | 0.0 (0) | 0.0 (0) |
|  | Adult weighing scale | 100 (10) | 0.0 (0) | 0.0(0) |
|  | Stadiometer(height equipment) | 100(10) | 0.0 (0) | ( )0.0(0) |
|  | Cholesterol | 0.0 (0) | 0.0 (0) | 100.0 (12) |

**Table 3:** Availability of selected medicines at the primary facilities(N=10)

| Condition | Drug | Consistent stock %<br>within the last three<br>months (n) | Inconsistent stock %<br>within the last three<br>months (n) | Never available<br>% (n) |
| --- | --- | --- | --- | --- |
| **HIV** | 1<sup>st</sup> Line ART | 100(10) | 0.0(0) | 0.0(0) |
|  | 2<sup>nd</sup> Line ART | 20.0(2) | 40.0(4) | 40.0 (4) |
|  | 3<sup>rd</sup> Line ART | 10.0(1) | 20.0 (2) | 70.0 (7) |
| **Hypertension** | Thiazide | 90.0(9) | 10.0(1) | 0.0(0) |
|  | Calcium Channel Blockers | 60.0(6) | 20.0(2) | 20.0(2) |
|  | ACE Inhibitors | 40.0(4) | 30.0(3) | 30.0(3) |
|  | Beta Blockers | 20.0(2) | 80.0(8) | 0.0(0) |
| **Diabetes mellitus** | Glibenclamide | 60.0(6) | 30.0(3) | 10.0(1) |
|  | Metformin | 50.0(5) | 50.0(5) | 0.0(0) |
|  | Dextrose | 60.0(6) | 40.0(4) | 0.0(0) |
|  | Glucose Injectable 10% Or 50% | 90.0(9) | 10.0(1) | 0.0(0) |
| **Chronic kidney disease** | Furosemide | 30.0(3) | 60.0(6) | 10.0(1) |
|  | Calcium Carbonate | 0.0(0) | 0.0(0) | 100(10) |
|  | Ferrous Sulphate | 70.0(7) | 10.0(1) | 10.0(1) |
|  | Vitamin D | 0.0(0) | 0.0(0) | 100(10) |

Participants reported inconsistent stock is sometimes due to low supply at the facilities as opposed to a lack of supply.

“*We don’t have glibenclamide, most of the time we usually have metformin. When it comes to hypertension most times we don’t have atenolol, amlodipine is usually few in stock, and we have hydrochlorothiazide. Frusemide is not enough. Most of the time, medication is there but in small quantity” (BT_mw08,female,medical assistant)*

Poor availability of medication affects patients who sometimes end up going home without filling their prescriptions.

*“For hypertension, it can be easier for those who are on HCTZ (hydrochlorothiazide) because they are the ones that are available …but those who are getting a combination of amlodipine and the others, they struggle because their medicine is not there” (Cz_mw12,male,medical assistant)*

The lack of essential medication also leads to unnecessary referrals to higher facilities which ends up being expensive for the patients and congests the systems. Participants believe consistent stock can reduce this problem.

*“We want to be given a full package of drugs so that when we say integrated NCD clinic, we should have a full package of medicine, other drugs like prinivil are not found here and for a patient with hypertension and diabetes, they feel better when such a drug is included in their dosage, but we do not have them here. Referring them is a waste of resources.” (BT_mw01,male, medical assistant)*

### Patient treatment and follow-up

Some participants acknowledged the need to manage multimorbidity because of potential drug interactions and disease complications.

*“if we do not help them with this problem urgently, it will also cause other problems…if we cannot manage, one (condition) can worsen another” (BT_mw06,female,nurse)”*

Integrated NCDs clinics were offered at half of the facilities (5/10) but were often limited to diabetes and hypertension, with other chronic NCDs being managed as general outpatients within the facility. Two facilities (2/10) reported providing integrated chronic disease management within ART clinics because the HIV guidelines provide for this (Figure 3).

**Figure 3:**
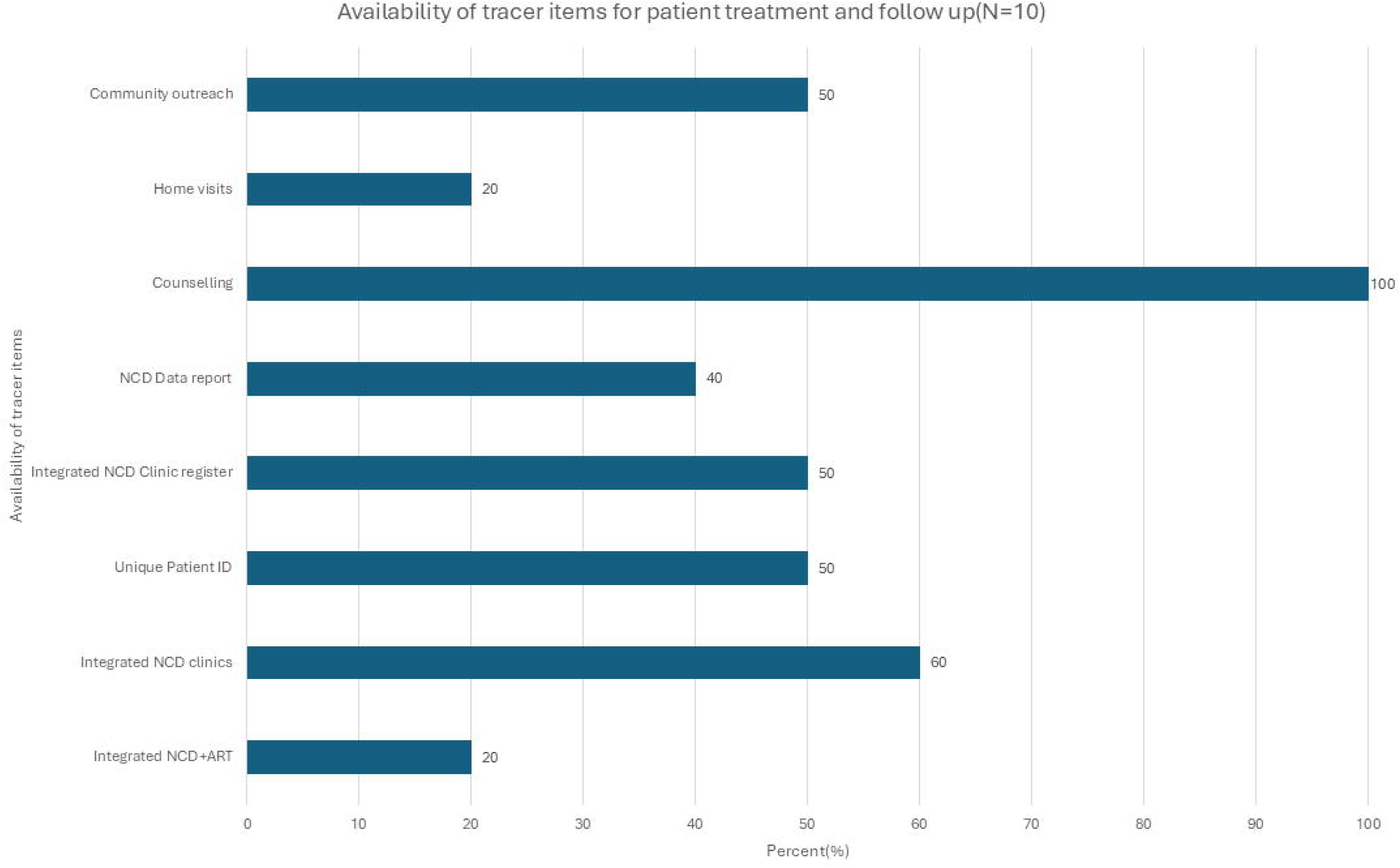
Availability of tracer item for patient treatment and follow-up domain.

“*They are assisted there at the ART clinic and they are not sent to the OPD again. They just come to get medication but they are seen right there in the clinic. We meet those patients often. That’s why the current HIV guidelines for those over 40 who are also on 13A (*Dolutegravir+Lamivudine+Tenofovir*) need to have their sugar and blood pressure checked routinely. It’s there in the guidelines.” (CZ_mw10,male,medical assistant)*

All participants reported providing counselling services, lifestyle modification and drug adherence advice in liaison with HSAs as part of pre-clinic health talks. This was limited to verbal conversations without any leaflets to take home.

*“We teach them on lifestyle and medication. We work closely with the HSAs who also give health talks on diet and lifestyle. Moving forward though, I would want to write them down and give them …leaflets or hard copy notes. “ (BT_mw05,female,medical assistant)*

*“…counselling is just verbal. Even the counselling is usually just general health education with HSAs, not one to one counselling” (BT_mw03,male,clinical officer)*

However, integration and a holistic approach to patient management were not always possible due to a lack of resources which led to prioritising some conditions over others.

*“We are supposed to have a team of nurses, HSAs, data clerks, and clinicians, but most of the time there are only HSAs and a clinician and no nurses, so it is very rare for us to read vital signs, maybe we just check the weight and only focus on nutrition.”* (BT_mw01,male,medical assistant)

All study facilities had outpatient registers and HIV patient IDs, half (5/5)provided unique patient ID numbers for NCDs allowing patient follow-up. HIV reports and audits were reported to the routinely produced in all facilities that provided HIV services while only 50.0% (5/10) of facilities provided NCD registers for new cases or recurring patients and only 40.0%(4/10) produced NCD reports. Despite keeping records of NCD patients, we were unable to visually verify data on disease burden at the time of data collection and none of the facilities produced an integrated chronic disease report.

## DISCUSSION

We examined service-specific availability and the readiness of twelve primary health facilities in Blantyre and Chiradzulu to manage patients with multimorbidity. Our findings shows some disparities in the availability for our selected tracer items as only 25%(3/12) of the surveyed facilities were “ready” to provide integrated care for chronic kidney disease, type 2 diabetes, HIV and hypertension. Most of the facilities failed to reach the minimum threshold for diagnostic equipment, essential medicines and treatment and follow-up.

Non-functional or unavailable diagnostic tools can be a major factor in delayed diagnoses, especially for people living with multimorbidity. Our data shows that most diagnostic equipment was either not available or available but non-functional. For example, we found limited availability of creatinine testing for chronic kidney disease which is contrary to the 2021 WHO PEN Plus operational plan for Malawi which indicated chronic kidney disease may be screened for at the primary facility, but management should be referred to secondary facilities.(21) Only 25%(4/12) of our facilities had urine glucose test-kits compared to similar low-income settings, such as 81% in Nepal and 87% in Afghanistan.(22) Our findings are like Ahmed et al. who analysed data from the 2019 Malawi Harmonised Health Facility Assessment and found fewer than 40% of PHCs were ready to manage 14 NCDs mostly due to a lack of diagnostic tools and medicines.(16) In Ethiopia, a high percentage of non-functional diagnostic equipment compromised chronic disease, which can affect multimorbidity management.(23)

Our findings are consistent with prior studies from Malawi which found that primary healthcare workers were not adequately trained to manage diabetes, hypertension and their complications.(24) Despite training being reported by our participants, they recommended even more trained staff as they currently rely on task shifting to HSAs. Inadequate number of trained human resources has been documented to compromise care.(25, 26) In our study, task shifting to HSAs reduced the burden of inadequate staff, as was observed in other studies conducted in similar settings.(27-29)

Similar to findings from Shiroya et al in 2021, we found a less than 70% availability of essential medicines to provide integrated care for selected NCDs and HIV. (30) However, when we examined the availability of medication for individual conditions, we found a stable supply of ARTs and varying availability of medication for diabetes and hypertension.(26)We show a better availability of a better availability of thiazides for hypertension(90%) and consistent findings for metformin diabetes (50%), compared to 64% and 48% respectively in 2014.(16) Low availability of type 2 diabetes medication has been noted in other low-income settings.(31, 32)We concur with the arguments raised by Ammoun et al, that the availability of trained staff and the availability of medication should go together, especially for patients living with multimorbidity.(33)

Record keeping was poor among our study facilities, with only half (n=5) of our facilities having NCD clinic registers, with even fewer facilities producing periodic NCD data reports. Our findings are worse than findings from Banda et al. in 2019, who found 91% of primary facilities in Malawi did not have NCD registers.(17) Poor NCD records have been reported to hinder the monitoring and evaluation of services for NCDs in other low-income countries.(34-36) Mulupi et al. noted that lack of NCD data impacted budget allocation for NCDs which in turn affects the availability of resources and NCD prioritisation. We believe the lack of NCD registers and reports (unlike the available HIV reports) reflects a lack of harmonisation within protocols and policies for reporting, compared to HIV.

### Strengths and limitations

Combining surveys and interviews provided a deeper and contextual understanding of the data. Our study highlights gaps in the implementation of integrated care recommended in the Malawi Health Sector Strategic Plan III (HSSP III) exemplified by inconsistency in stock for medication, inadequate availability of diagnostic equipment. This study highlights the importance of adequate resource allocation to support policy implementation.

Our study has some limitations. Firstly, our findings are limited to a sample size of 12 public funded primary facilities which may not be generalisable to all primary health facilities in Malawi. We did not calculate differences in facility readiness based on facility characteristics such as location and district, as this was beyond the scope of our study. Secondly, our focus has been on four conditions and did not include chronic hepatitis, chronic respiratory diseases, cancers and mental health, which can all be possible multimorbidity.

Thirdly, the cross-sectional nature of our study may not reflect readiness throughout the year, although we are aware that stockouts tend to be seasonal and not steady. The survey may oversimplify the diagnosis process of conditions that require several readings such as hypertension and type 2 diabetes. The tool we used does not fully capture the quality of patient management, such as monitoring drug-drug interactions or changing prescriptions in light of side effects. The cutoff point of 70%, although recommended by the WHO, is still discretionary and the absolute binary outcome of “ready” or “not ready” does not reflect the quality of available services.

Further research is required to assess the availability and readiness of both for-profit and not-for-profit primary health facilities. A comparison of district performance may also be necessary to determine district-level differences in the face of decentralised health management.

### Conclusion

The primary health care in Malawi does not meet the minimum readiness score to provide integrated chronic disease care for common multimorbidity at primary health level in Blantyre and Chiradzulu. This undermines a decentralised approach, creating congestion at higher levels of the systems and leading to opportunities for early diagnosis and prevention of complications or hospital admission. The gaps in providing integrated chronic disease care in outpatient clinics may be attributed to the disproportionate impact of poor supply of diagnostic equipment and frequent drug stock-outs. The presence of trained staff and guidelines in communities means there is potential to use task shifting to deliver screening, preventive and follow-up care at community level, but this can only happen with functional diagnostic equipment and drugs consistently in stock. Emphasis on periodic quality assurance and combined multimorbidity audits could further highlight the gaps in service delivery and give clues to improvement.

## Declarations

### Ethical approval and Consent to participate

The study was conducted according to the guidelines of the Declaration of Helsinki. The College Medicine Research & Ethics Committee (Ref P.11/21/3462) (Malawi) and the Liverpool School of Tropical Medicine (Ref. 21-086) (UK) gave ethical approval for this study. All participants provided written consent to be included in the study prior to data collection. Participants were assured the right to withdraw from the study at any time without needing to justify their decision. Participants also gave written consent for their anonymised quotes using participant identification codes, to be published.

### Consent for publication

Not applicable

### Author contributions

Conceptualisation: GTB, FL, JR, RM, MT

Data collection: GTB

Formal data analysis: GTB

Supervision: FL, RM, MT

Drafting manuscript: GTB

Review and editing: All authors

Funding acquisition: FL,JR,AC,JH,ASM,MN,MR,FS,HS,MT,SU,MSC,EW

### Acknowledgements

We would like to acknowledge the management and staff at our study sites for their support during data collection. We would also like that acknowledge the works and contributions of the members of the Multilink consortium. The full list of the multilink members can be found in Additional file 4.

### Data availability

Data is provided within the manuscript or supplementary information files. Any additional data are, however, available from the authors upon reasonable request and with the permission of the Multilink Consortium.

