## Additional file 2 for "Service availability and readiness for out-patient integrated management of multimorbidity in public primary health facilities in Malawi: A mixed methods analysis"

**Additional file 2: Tracer indicators for service availability and readiness**

| **Domain** | Resources |
|---|---|
| Basic amenities | •Room for audio and visual privacy<br>•Communication device<br>•Access to a computer<br>•Emergency transportation |
| Staff training and guidelines | •Trained staff on cardiovascular disease diagnosis and treatment<br>• Trained staff on diabetes diagnosis and treatment<br>• Trained staff on HIV diagnosis and treatment<br>• Clinical guidelines for the management of HIV<br>•Malawi Standard treatment guidelines<br>•WHO PEN protocol<br>•Wall chart on NCD screening |

| | |
|---|---|
| Basic diagnostic equipment | • Blood pressure apparatus<br>• Weighing machine<br>• Stadiometer<br>• Stethoscope<br>• Blood glucose test<br>• Urine dipstick-glucose<br>• Microscopy urinalysis<br>• Creatinine<br>• HIV diagnostic capacity<br>• CD4 count<br>• PCR for viral load<br>• Cholesterol |
| Essential Medicines | • ACE inhibitor (enalapril)<br>• Thiazide<br>• Beta-blocker (atenolol)<br>• Calcium channel blocker (amlodipine)<br>• Metformin<br>• Glibenclamide |

| | |
|---|---|
| | • Glucose injectable solution 10% or 50%<br>• Dextrose 5%<br>• Iron<br>• Ferrous sulphate<br>• Calcium carbonate<br>• Vitamin D supplement<br>• Furosemide<br>• Aspirin (acetylsalicylic acid) capsules/tablets<br>•ARTs standard for patients 30kgs+<br>•1st line ARTs<br>• 2nd line ARTs<br>• 3rd like ARTs |
| Patient treatment and follow-up | • Outpatient registers<br>• Out-patient clinics<br>• Data reports<br>• Counselling services on lifestyle risk factors<br>• Self-management counselling<br>•Patient identification numbers |

| | |
|---|---|
| | •Community outreach |
