## Additional file 3 for "Service availability and readiness for out-patient integrated management of multimorbidity in public primary health facilities in Malawi: A mixed methods analysis"

Additional file 3: Readiness index by facility out of 44 tracer items. Vertical black line indicates cutoff (70%)

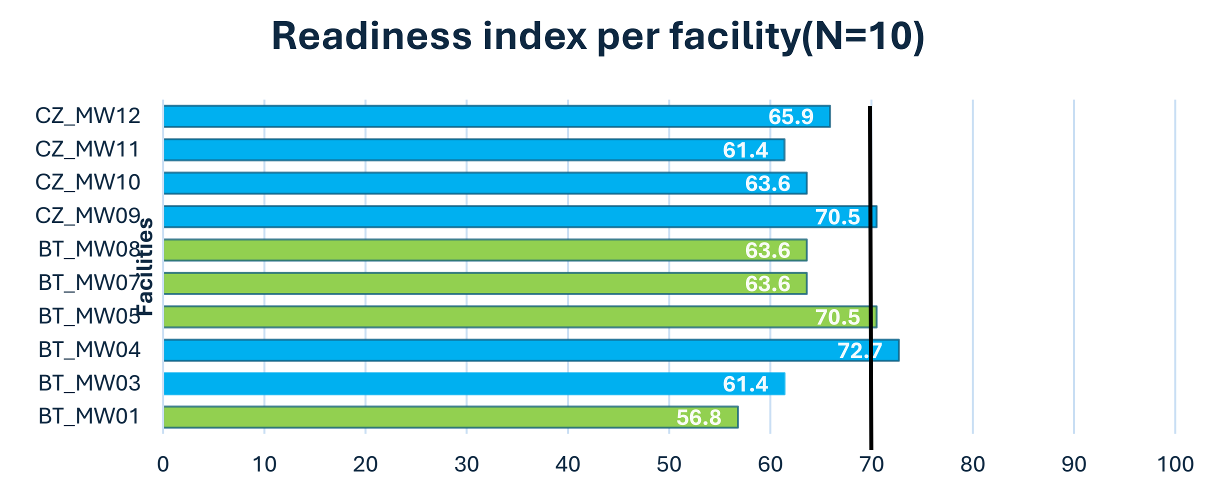


| | BT_MW 01 | BT_MW 03 | BT_MW 04 | BT_MW 05 | BT_MW 07 | BT_MW 08 | CZ_MW 09 | CZ_MW 10 | CZ_MW 11 | CZ_MW 12 |
|---|---|---|---|---|---|---|---|---|---|---|
| Percentage | 56.8 | 61.4 | 72.7 | 70.5 | 63.6 | 63.6 | 70.5 | 63.6 | 61.4 | 65.9 |
| | 25 | 27 | 32 | 31 | 28 | 28 | 31 | 28 | 27 | 29 |

**Mean percentage of available healthcare services to diagnose and manage multimorbidity**

Percentage
