## Additional file 4 for "Service availability and readiness for out-patient integrated management of multimorbidity in public primary health facilities in Malawi: A mixed methods analysis"

**Members of the Multilink Consortium**

Felix Limbani(funding)

Jamie Rylance(funding)

Augustine Choko(funding)

Paul Dark(funding)

Marc Henrion

Julian Hertz(funding)

Ben Morton

Adamson Muula(funding)

Mulinda Nyirenda(funding)

Matthew Rubach(funding)

Charity Salima(funding)

Francis Sakita(funding)

Hendry Sawe(funding)

Stephen Spencer

Miriam Taegtmeyer(funding)

Sarah Urasa(funding)

Sarah White(funding)

Alice Rutta

Gimbo Hyuha(funding)

Ibrahim Simiyu

Sangwani Salimu

Gift Treighcy Banda

Nateiya Yongolo

Rhona Mijumbi

Jacob Phulusa

Stephen Gordon

Alice Rutta

Blandina Mmbaga

Rachel Mangoni

Robert Chuwa

Sanjura Biswaro

Albert Mukatipa

Alfred Muyaya

Bright Mnesa

Amy Smith

Cathy Waldron

Ewan Tomeny

Laura Rosu

Nicola Desmond

Deborah Nyirenda

Marlen Chawani(funding)

Juma Mfinanga

Catherine Wu

Kathy Rowan

Constantine Tarimo

Frank Kimaro

Mwamini Kacheuka

Rose Freddy

Yesse Bumija

Zanuni Kweka

Katie Davies

Ranjeeta Thomas

Rashida Ferrand

Jamie Rylance

Charity Rylance

Diana Msindira

Genesis Msindira

Asimenye Kayuni

Kate Mangulama

Chiku Simbano

Frank Gugu

Hussein Moremi

Noela Mpili

Nsajigwa Mwakyambiki

Nuhu Richard

Ramadhani Mashoka

Vicky Mlele

Yusuph Chimpaye

Abdulaziz Abdallah Nassoro

Benjamin Paulo Mwenda

Grasiana Kimario

Herieth Cliff Mushi

Naftari Mahimbo

Safina Baleche

Joanna Jozefiak

Yusuf Iqbal

Jane Goudge

Arianna Braccioni

Lucy Keyala

Beatrice Chinoko

Slyvester Kaimba

Mercy Mkandawire

Peter Mandala

Maureen Kandiero

Matthew Mlongoti

Treighcy Gift Banda

Rhona Majumbi

Mullinda Nyirenda

Francis Sakita

Sarah Urasa

Blandina Mmbaga

Frank Michael Kimaro

Robert Chuwa

Zanuni Kweka

Sanjura Biswaro

Mwamini Kacheuka

Yesse Bumija

Martha Oshoseni

Gideon Tesha

Herieth Cliff Mushi

Hussein R. Moremi

Eve Worrall(funding)
