## Additional file 1 for "Service availability and readiness for out-patient integrated management of multimorbidity in public primary health facilities in Malawi: A mixed methods analysis"

Service Availability Assessment tool adapted from the WHO PEN, PEN- PLUS, MSPA, and WHO 2021 Essential Medicines list

**Name of facility………………………………………………….. Date of visit (DD/MM/YY)………/………/………..**
**Location of the facility (km from Referral hospital) …………………………………………………..**
**District/City…………………………………………………………………………**
**Facility density in district……………………………………………………………………..**
**Urban/rural…………………………………………………….**
**Facility opening hours…………………………………………….  Total Hours……………..**
**Number of Out-patient adults seen in the last calendar month…………………………… /NOT KNOWN**
**Number of adults in the clinic catchment area…………………………………………………..**
**Are there user-fee charges for clients? YES/NO**

| | **Code** | **Item** | **Available YES NO** | | **Functional YES NO** | |
|---|---|---|---|---|---|---|
| **Basic Amenities** | 001 | Room for auditory and visual privacy for consultation | 1 | 2 | 1 | 2 |
| | 001A | Communication equipment (phone or short-wave radio) | 1 | 2 | 1 | 2 |
| | 001B | Access to a computer | 1 | 2 | 1 | 2 |
| | 001C | Emergency transportation | 1 | 2 | 1 | 2 |
| **Basic equipment** | 002 | Thermometer | 1 | 2 | 1 | 2 |
| | 002A | Stethoscope | 1 | 2 | 1 | 2 |
| | 002B | Blood pressure apparatus | 1 | 2 | 1 | 2 |
| | 002C | Glucometer | 1 | 2 | 1 | 2 |
| | 002D | HIV Diagnostic capacity | 1 | 2 | 1 | 2 |
| | 002E | Adult weighting scale | 1 | 2 | 1 | 2 |
| | 002F | Adult height measuring equipment | 1 | 2 | 1 | 2 |
| **Diagnostic capacity** | | | | | | |
| | 003 | Urine dipstick glucose | 1 | 2 | 1 | 2 |
| | 003A | Microscopy urinalysis | 1 | 2 | 1 | 2 |
| | 003B | CD4 count | 1 | 2 | 1 | 2 |
| | 003C | PCR for viral load | 1 | 2 | 1 | 2 |
| | 003D | Creatinine | 1 | 2 | 1 | 2 |

Approved by
College of Medicine
18-Aug-2023
(COMREC)
Research and Ethics Committee

| | | | | | | |
|---|---|---|---|---|---|---|
| | 003E | Cholesterol | 1 | 2 | 1 | 2 |
| | 003F | ECG machine | 1 | 2 | 1 | 2 |
| | 003G | Copy of Clinical guidelines for HIV present at facility | 1 | 2 | 1 | 2 |
| **Clinical guidelines for NCD present at the facility** | | | **Available** | | **Not Available** | **Not sure** |
| | 004 | WHO PEN protocol | 1 | | 2 | 9 |
| | 004A | Wall charts on NDC screening | 1 | | 2 | 9 |
| | 004B | Malawi standard treatment guidelines | | | | 9 |
| | 004C | Wall chart of NCD screening | 1 | | 2 | 9 |
| **Patient care** | | | | | | |
| | 005 | Separate NCD clinic | 1 | | 2 | |
| | 005A | Do patients have a unique ID | 1 | | 2 | |
| | 005B | Is there a facility register for patients seen? | 1 | | 2 | |
| | 005C | Is there a separate NCD register? | 1 | | 2 | |
| | 005D | Are there NCD data reports produced? | 1 | | 2 | |
| **Services available** | | | | | | |
| | 006 | Patient education on lifestyle and nutrition | 1 | | 2 | |
| | 006A | Patient counselling for self-management | 1 | | 2 | |
| | 006B | Family counselling | 1 | | 2 | |
| | 006C | Home visits for patients | 1 | | 2 | |
| | 006D | Community engagement for multimorbidity activities | 1 | | 2 | |
| **Staff available** | 007 | Nurses with BSc | 1 | | 2 | |
| | 007A | Nurses with a Dip | 1 | | 2 | |
| | 007B | Clinician Dip | 1 | | 2 | |
| | 007C | Clinician BSc | 1 | | 2 | |
| | 007D | Medical Assistant | 1 | | 2 | |
| | 007E | Pharmacist | 1 | | 2 | |
| | 007F | Pharmacy Technologist | 1 | | 2 | |
| | 007G | Rehabilitation Officer | 1 | | 2 | |
| | 007H | Rehabilitation Technician | 1 | | 2 | |
| | 007I | Health Surveillance assistant | 1 | | 2 | |
| **Trained staff** | 008A | Are there staff dedicated to NCDs? | 1 | | 2 | |

Approved by
College of Medicine
18-Aug-2023
(COMREC)
Research and Ethics Committee

| | | | | | | |
|---|---|---|---|---|---|---|
| | 008B | Are there staff dedicated to HIV services? | 1 | 2 | | |
| | 008C | Are there staff members that received pre-service training to diagnose and treat chronic conditions? | 1 | 2 | | |
| | 008D | Do you have access to CPDs on Chronic disease management at the facility? | 1 | 2 | | |
| | 008E | Were they trained to diagnose and prescribe for NCDs | 1 | 2 | | |
| | 008F | Were they trained to monitor and look out for drug interaction? | 1 | 2 | | |
| **Essential Medicines** | | | | | | |
| | | **Medicine** | **Available** | **Not Available** | **Never available** | **Consistent stock for the last 3 months** |
| **Cardiovascular** | 009 | Glyceryl trinitrate | 1 | 2 | 9 | 1 |
| | 009A | Verapamil | 1 | 2 | 9 | 1 |
| | 009B | Furosemide (Bumetanide, Torasemide) | 1 | 2 | 9 | 1 |
| | 009C | Aspirin Capsule/tablet | 1 | 2 | 9 | 1 |
| **Hypertension** | 010 | Beta-blocker (atenolol, metoprolol, bisoprolol, carvedilol) | 1 | 2 | 9 | 1 |
| | 010A | ACE inhibitor (Enalapril*, Lisinopril, Ramipril, Perindopril) | 1 | 2 | 9 | 1 |
| | 010B | calcium channel blocker (Amlodipine tablet or an alternative) | 1 | 2 | 9 | 1 |
| | 010C | Thiazide (Hydrochlorothiazide)* | 1 | 2 | 9 | 1 |
| | 010D | Lisinopril + Amlodipine | 1 | 2 | 9 | 1 |
| | 010E | Lisinopril + Hydrochorothiazide | 1 | 2 | 9 | 1 |
| | 010F | Telmisartan + Amlodipine | 1 | 2 | 9 | 1 |
| | 010G | ARB (Telmisartan, Losartan,) | 1 | 2 | 9 | 1 |

Approved by
College of Medicine
18-Aug-2023
(COMREC)
Research and Ethics Committee

| | | | | | | |
|---|---|---|---|---|---|---|
| **Diabetes Mellitus** | | | | | | |
| | 011A | Glibenclamide tablet | 1 | 2 | 9 | 1 |
| | 011B | Glucose Injectable solution 10% or 50% | 1 | 2 | 9 | 1 |
| | 011C | Insulin regular injection | 1 | 2 | 9 | 1 |
| | 011D | Metformin tablet | 1 | 2 | 9 | 1 |
| | 011E | Glucose injectable 5% dextrose | 1 | 2 | 9 | 1 |
| **Chronic Kidney disease** | 012 | Intravenous Iron | 1 | 2 | 9 | 1 |
| | 012A | Ferrous salt (sulphate) | 1 | 2 | 9 | 1 |
| | 012B | Sodium glucose co transporter 2 inhibitors i.e canagliflozin, dapagliflozin, and empagliflizin | 1 | 2 | 9 | 1 |
| | 012C | Calcium carbonate | 1 | 2 | 9 | 1 |
| | 012D | Vitamin D supplements | 1 | 2 | 9 | 1 |
| | 012E | Erythropoietin | 1 | 2 | 9 | 1 |
| **HIV** | | | | | | |
| Standard for all patients 30 kg+ | 013 | Dolutegravir+ Lamivudine + Tenofovir (DTG + 3TC + TDF) **13A** | 1 | 2 | 9 | 1 |
| | 013A | Abacavir + Lamivudine + Tenofovir (ABC +3TC+ TDF) **15A** | 1 | 2 | 9 | 1 |
| Alternative if (relative) DTG contraindications | 013B | Efivarenz +Lamivudine +Tenofovir (EFV+ 3TC + TDF) **5A** | 1 | 2 | 9 | 1 |
| 1st line drugs not used for ART initiation | 013C | Zidovudine+ Lamivudine+ Efivarenz (AZT+ 3TC+EFV) **4A** | 1 | 2 | 9 | 1 |
| | 013D | Abacavir+ Lamivudine+ lopinavir/ritonavir (ABC+ 3TC+ LPV/r)**9A** | 1 | 2 | 9 | 1 |
| | 013E | Zidovudine + Lamivudine+ Dolutegravir(AZT + 3TC+ DTG) **14A** | 1 | 2 | 9 | 1 |
| | 013F | Abacavir+Lamivudine+ Efivarenz (ABC+3TC+EFV) **17A** | 1 | 2 | 9 | 1 |
| 2nd line ARTs | 13G | Tenofovir+ Lamivudine+ Atazanivir (TDF+3TC+ATV)**7A** | 1 | 2 | 9 | 1 |
| | 013H | Zidovudine +Lamivudine+Atazanir/ritonavir(AZT+3TC+ATV/r)**8A** | 1 | 2 | 9 | 1 |



Approved by
College of Medicine
18-Aug-2023
(COMREC)
Research and Ethics Committee

| | | | | | | |
|---|---|---|---|---|---|---|
| | 013I | Tenofovir + Lamivudine + Lopinavir/ritonavir (TDF+3TC+LPV/r)**10A** | 1 | 2 | 9 | 1 |
| | 013J | Zidovudine+Lamivudine+Lopinavir(AZT+ 3TC+LPV)**11A** | 1 | 2 | 9 | 1 |
| 3rd line ARTs | 013K | Darunavir+ritonavir+Dolutegariv(DRV+r+DTG)**12A** | 1 | 2 | 9 | 1 |



Approved by
College of Medicine
18-Aug-2023
(COMREC)
Research and Ethics Committee
